# Association between benzodiazepine receptor agonist use and increased mortality among patients hospitalized for COVID-19: results from an observational study

**DOI:** 10.1101/2021.02.18.21252004

**Authors:** N. Hoertel, M. Sánchez-Rico, E. Gulbins, J. Kornhuber, R. Vernet, N. Beeker, A. Neuraz, C. Blanco, M. Olfson, G. Airagnes, C. Lemogne, J. M. Alvarado, M. Arnaout, P. Meneton, F. Limosin, >On behalf of AP-HP / Universities / INSERM COVID-19 research collaboration and AP-HP COVID CDR Initiative

## Abstract

**Objective:** To examine the association between benzodiazepine receptor agonist (BZRA) use and mortality in patients hospitalized for COVID-19.

**Methods:** We conducted an observational multicenter retrospective cohort study at AP-HP Greater Paris University hospitals. The sample involved 14,381 adult patients hospitalized for COVID-19. 686 (4.8%) inpatients received a BZRA within at the time of hospital admission at a mean daily diazepam-equivalent dose of 19.7 mg (SD=25.4). The study baseline was the date of hospital admission and the primary endpoint was death. We compared this endpoint between patients who received BZRAs and those who did not in time-to-event analyses adjusted for patient characteristics (such as age, sex, obesity and comorbidity) and other medications. The primary analysis was a Cox regression model with inverse probability weighting (IPW).

**Results:** Over a mean follow-up of 14.5 days (SD=18.1), the primary endpoint occurred in 186 patients (27.1%) who received a BZRA and in 1,134 patients (8.3%) who did not. There was a significant association between BZRA use and increased mortality both in the crude analysis (HR=3.20; 95% CI=2.74-3.74; p<0.01) and in the primary IPW analysis (HR=1.61; 95% CI=1.31-1.98, p<0.01), with a significant dose-dependent relationship (HR=1.55; 95% CI=1.08-2.22; p=0.02). This association remained significant in multiple sensitivity analyses.

**Conclusions:** BZRA use was associated with increased mortality among patients hospitalized for COVID-19 with a dose-dependent relationship, suggesting a potential benefit of decreasing dose or tapering off these medications when possible in these patients.

## 1. Introduction

Global spread of the novel coronavirus SARS-CoV-2, the causative agent of coronavirus disease 2019 (COVID-19), has created an unprecedented infectious disease crisis worldwide (1,2). Benzodiazepine receptor agonists (BZRAs), including benzodiazepines and Z-drugs, potentiate the rapid neuroinhibitory effect of the neurotransmitter gamma-aminobutyric acid (GABA) in the brain and spinal cord. These medications are commonly prescribed for anxiety and sleep problems (3) and as muscle relaxants and pre-medications in anesthesia, but are also effective for treating epilepsy, alcohol withdrawal syndrome, and acute behavioral disturbance (4). Potential deleterious effects associated with these medications are well documented. Several studies (5–11), although not all (12–15), have found a significant association between BZRAs and increased all-cause mortality. BZRAs have also been associated with increased risk of infection (16), including pneumonia (17), asthma exacerbation (18) and respiratory depression (19).

To our knowledge, there are no data on the association of BZRA use with mortality among patients hospitalized for COVID-19. Following a recent release by the US Food and Drug Administration (FDA) of a drug safety communication warning patients and health care providers about the risks of BZRA use (20), it is important to investigate whether these medications are associated with increased mortality among patients with COVID-19. If this were the case, a second unanswered question is whether all BZRAs or specific BZRA treatments are associated with this risk. This knowledge might help guide clinicians on the choice of medications for patients with COVID-19 with clinical indications for BZRAs.

In this report, we examined the association between BZRAs and mortality in the Assistance Publique-Hôpitaux de Paris (AP-HP) Health Data Warehouse, which collects data on all hospitalizations for COVID-19 in Greater Paris University hospitals since the beginning of the epidemic. If a significant association were found, we sought to perform additional exploratory analyses to examine whether this association was specific to all or certain BZRA treatments. Our primary hypothesis was that BZRA use would be associated with increased mortality among patients hospitalized for COVID-19 in time-to-event analyses adjusting for patient characteristics (such as age, sex, obesity and comorbidity) and other medications (other psychotropic medications and medications prescribed according to compassionate use or as part of a clinical trial). If this were the case, our secondary hypotheses were that this association would be dose-dependent with higher doses conferring greater risk, and partially explained by respiratory depression.

## 2. Methods

### 2.1. Setting and Cohort Assembly

We conducted a multicenter observational retrospective cohort study at 36 AP-HP hospitals. We included all adults aged 18 years or over who have been admitted to these medical centers from the beginning of the epidemic in France, i.e. January 24^th^, until May 1^st^. COVID-19 was ascertained by a positive reverse-transcriptase–polymerase-chain-reaction (RT-PCR) test from analysis of nasopharyngeal or oropharyngeal swab specimens. This observational study using routinely collected data received approval from the Institutional Review Board of the AP-HP clinical data warehouse (decision CSE-20-20_COVID19, IRB00011591). AP-HP clinical Data Warehouse initiatives ensure patient information and informed consent regarding the different approved studies through a transparency portal in accordance with European Regulation on data protection and authorization n°1980120 from National Commission for Information Technology and Civil Liberties (CNIL). All procedures related to this work adhered to the ethical standards of the relevant national and institutional committees on human experimentation and with the Helsinki Declaration of 1975, as revised in 2008.

### 2.2. Data sources

We used data from the AP-HP Health Data Warehouse (‘Entrepôt de Données de Santé (EDS)’). This warehouse contains all available clinical data on all inpatient visits for COVID-19 to any Greater Paris University hospital. The data included patient demographic characteristics, vital signs, laboratory test and RT-PCR test results, medication administration data, medication lists during current and past hospitalizations in AP-HP hospitals, current diagnoses, discharge disposition, and death certificates.

### 2.3. Variables assessed

We obtained the following data for each patient at the time of the hospitalization through electronic health records (17,18): sex, age, hospital, obesity, self-reported current smoking status, any medical condition associated with increased clinical severity related to COVID-19 or BZRA use (21–24), any current psychiatric disorder, any medication prescribed according to compassionate use or as part of a clinical trial (25), any psychotropic medication, including any antidepressant (26), mood stabilizer (i.e. lithium or antiepileptic medications with mood stabilizing effects), and antipsychotic medication, respiratory depression, defined by a respiratory rate < 12 breaths/min or a resting peripheral capillary oxygen saturation in ambient air < 90% (25), and other clinical markers of disease severity, defined as having temperature > 40°C or systolic blood pressure < 100 mmHg or respiratory rate > 24 breaths/min or plasma lactate levels higher than 2 mmol/L (25). For these two later variables, a third category for missing data was added. These variables are detailed in **eMethods**.

### 2.4. Benzodiazepine receptor agonist (BZRA) use

Study baseline was defined as the date of hospital admission. BZRA use was defined as receiving these medications at baseline, i.e. within the first 48 hours of hospital admission, and before the end of the index hospitalization or death, to minimize potential confounding effects of late prescription of BZRAs, which can be used in palliative care. We used this delay because we considered that, in a context of overwhelmed hospital units during the COVID-19 peak incidence, all patients may not have received or been prescribed their usual medication regimens the first day of their hospital admission, or this treatment may not have been recorded at this time.

### 2.5. Primary endpoint

The primary endpoint was the time from study baseline to death. Patients without an end-point event had their data censored on May 1^st^, 2020.

### 2.6. Statistical analysis

We calculated frequencies of all baseline characteristics described above in patients receiving or not receiving BZRAs and compared them using standardized mean differences (SMD).

To examine the association between BZRA use and the endpoint of death, we performed Cox proportional-hazards regression models (27). To help account for the nonrandomized prescription of BZRAs and reduce the effects of confounding, the primary analysis used propensity score analysis with inverse probability weighting (IPW) (19,28). The individual propensities for receiving any BZRA were estimated using a multivariable logistic regression model that included patient characteristics (i.e., sex, age, hospital, obesity, current smoking status, any significant medical condition, and any current psychiatric disorder) and other other medications, including any medication prescribed according to compassionate use or as part of a clinical trial and other psychotropic medications, comprising any antidepressant, any mood stabilizer, and any antipsychotic medication (see **eMethods**). In the inverse-probability-weighted analyses, the predicted probabilities from the propensity-score models were used to calculate the stabilized inverse-probability-weighting weights (28). The association between BZRA use and the endpoint was then estimated using an IPW Cox regression model. In case of non-balanced covariates, an IPW multivariable Cox regression model adjusting for the non-balanced covariates was also performed. Kaplan-Meier curves were performed using the inverse-probability-weighting weights (29,30), and their pointwise 95% confidence intervals were estimated using the nonparametric bootstrap method (30).

We conducted five sensitivity analyses. First, we performed a multivariable Cox regression model including as covariates the same variables as in the IPW analysis. Second, we used a univariate Cox regression model in a matched analytic sample using a 1:1 ratio, based on the same variables used for the IPW analysis and the multivariable Cox regression analysis. To reduce the effects of confounding, optimal matching was used in order to obtain the smallest average absolute distance across all clinical characteristics between exposed patients and non-exposed matched controls. Third, to examine a potential indication bias of prescription of BZRAs in ICUs as a possible treatment for palliative care or as an aid to oral intubation, we reproduced the main analyses after excluding all patients who had been hospitalized in ICUs. Fourth, we examined whether our findings were similar in models imputing missing data using multiple imputation (31) instead of excluding patients with any missing data as done in the main analyses. Finally, to examine the potential influence on the results of excluding patients who received BZRA after 48 hours from hospital admission, we reproduced the main analysis while including all participants who received any BZRA at any time from hospital admission until the end of the index hospitalization or death, and considering BZRA use as a time-dependent variable (27). In that analysis, patients who received any BZRA after hospital admission enter the analysis risk-sets at the time of actual first initiation of the treatment. In all these analyses, weighted Cox regression model was used when proportional hazards assumption was not met (32).

We performed several additional exploratory analyses. First, we searched for a potential dose-dependent relationship by testing the association between daily dose of BZRA received at baseline (converted into diazepam-equivalent dose (33) and dichotomized at the mean value) and the endpoint, adjusted for the same covariates as used in the main analysis, among patients who received a BZRA. Second, we examined whether respiratory depression or other clinical markers of disease severity may at least partly explain this association by adjusting successively for these variables. Finally, we examined the relationships between each BZRA and mortality using the same statistical approach as described for the main analysis.

For all associations, we performed residual analyses to assess the fit of the data, check assumptions, including proportional hazards assumption using proportional hazards tests and diagnostics based on weighted residuals (27,34), and examined the potential influence of outliers. To improve the quality of result reporting, we followed the recommendations of The Strengthening the Reporting of Observational Studies in Epidemiology (STROBE) Initiative. Because our main hypothesis focused on the association between BZRA use and mortality, statistical significance was fixed *a priori* at two-sided p-value <0.05. Only if a significant association were found in the main analysis, we planned to perform additional exploratory analyses as described above. All analyses were conducted in R software version 2.4.3 (R Project for Statistical Computing).

## 3. Results

### 3.1. Characteristics of the cohort

Of the 17,131 patients with a positive COVID-19 RT-PCR test who had been hospitalized for COVID-19, 1,963 patients (11.5%) were excluded because of missing data or their young age (i.e. less than 18 years old of age). In addition, of the 1,473 patients who received a BZRA at any time during the visit, 787 were excluded because they were prescribed it more than 48 hours after hospital admission. Of the remaining 14,381 adult patients, 686 (4.8%) received a BZRA in the first 48 hours of hospitalization at a mean diazepam-equivalent dose of 19.7 mg per day (SD=25.4, median=10.0, lower quartile=5.0, higher quartile=22.9, range=1.85-240.0 mg) (**eFigure 1**). Of these 686 patients, 41.1% (N=282) had a prescription of BZRA during a prior hospitalization at AP-HP in the past 6 months, and 36.4% (N=183) received at least two BZRA medications. The median delay from hospital admission to first prescription of BZRA was 0.94 days (SD=0.55).

PT-PCR test results were obtained after a median delay of 1.2 days (SD=12.7) from hospital admission date. This median delay was of 1.0 day (SD=11.6) in the exposed group and of 1.2 day (SD=12.8) in the non-exposed group.

Over a mean follow-up of 14.5 days (SD=18.1; median=7 days), 1,320 patients (9.2%) had an end-point event at the time of data cutoff on May 1^st^. Among patients who received a BZRA, the mean follow-up was 12.8 days (SD=13.1, median=8 days), while it was of 14.5 days (SD=18.3, median=7 days) in those who did not.

All baseline characteristics were independently and significantly associated with the endpoint. A multivariable Cox regression model showed that age, sex, hospital, any medical condition, any current psychiatric disorder, respiratory depression and any other clinical marker of severity were significantly and independently associated with the endpoint (**eTable 1**).

The distributions of patient characteristics according to BZRA use are shown in **Table**

**1**. In the full sample, BZRA use significantly differed according to all characteristics except for sex ratio, and the direction of the associations indicated older age and greater medical severity of patients receiving any BZRA. After applying the propensity score weights, these differences were substantially reduced. In the matched analytic sample, there were no significant differences in any characteristic, except the greater prevalence of men in the non-exposed group (**Table 1**).

**Table 1.**
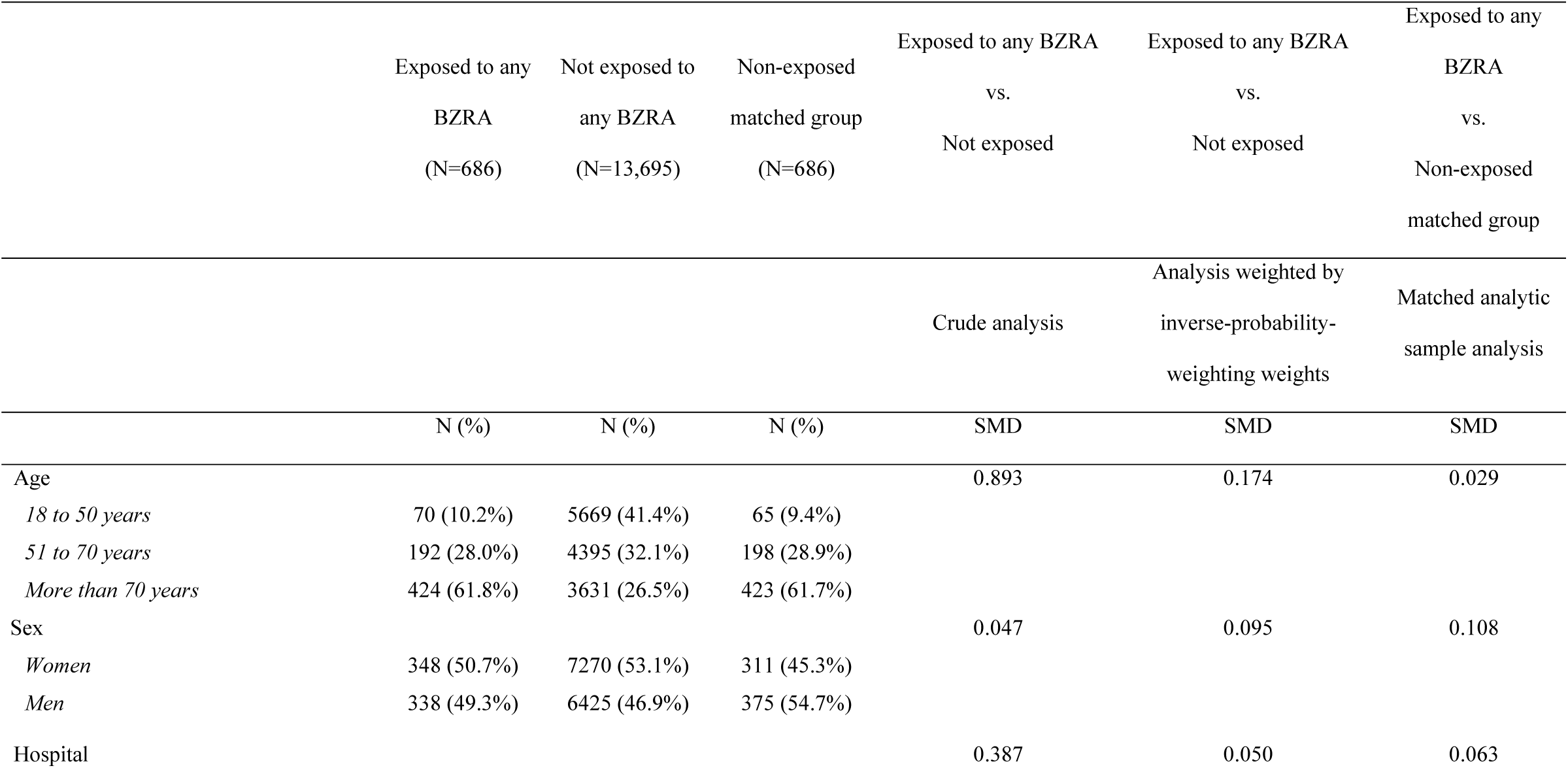
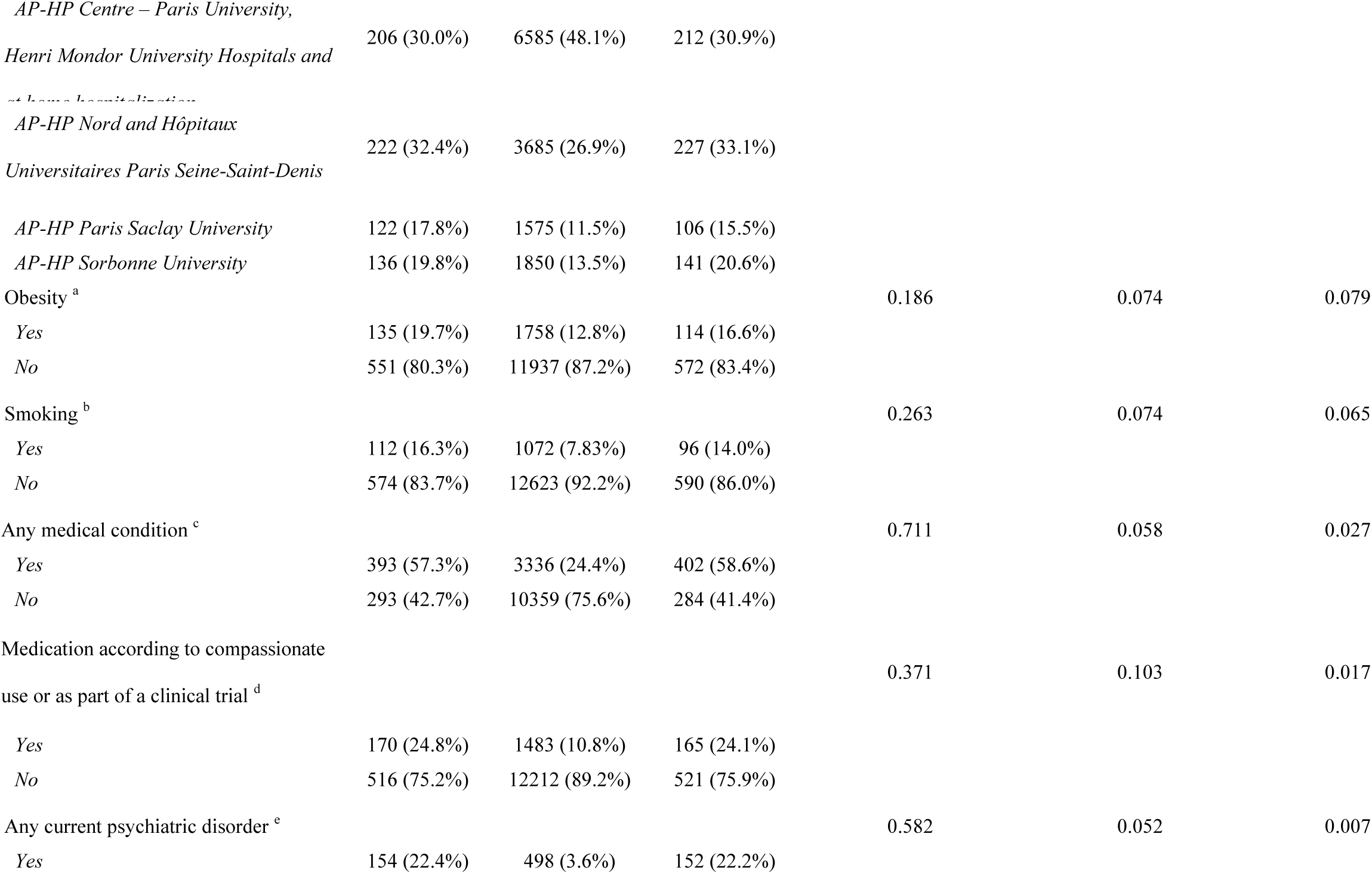
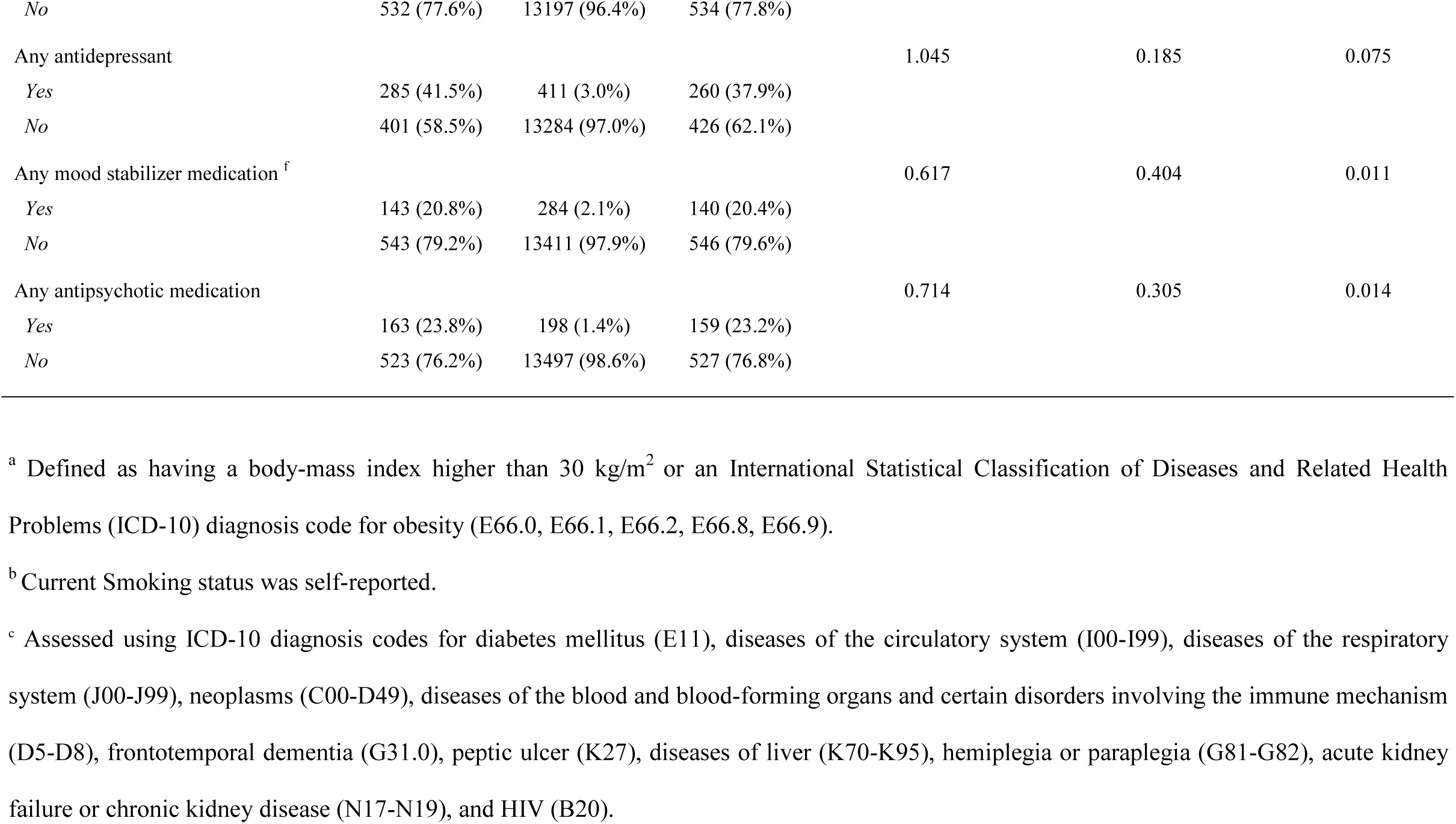
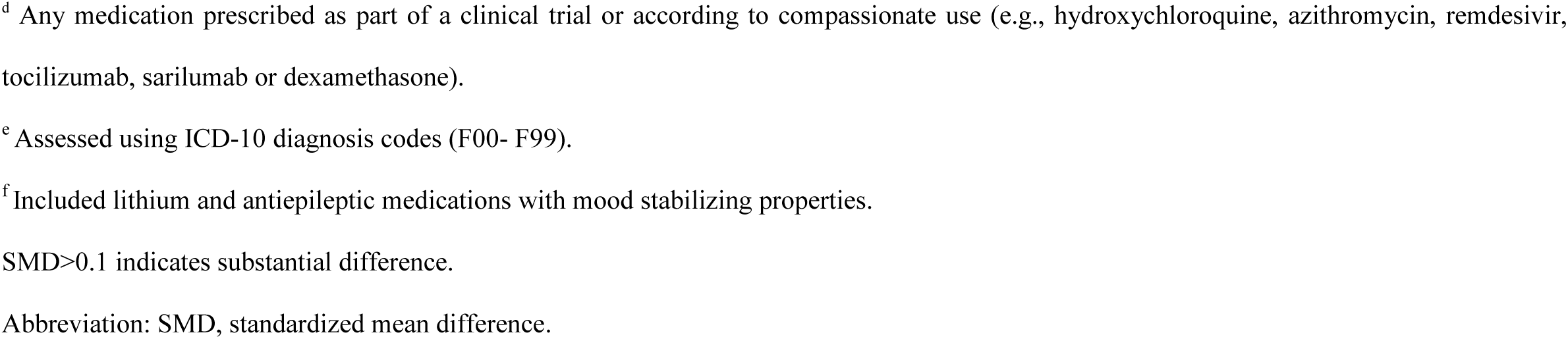
Characteristics of patients with COVID-19 receiving or not receiving a benzodiazepine receptor agonist (BZRA) at baseline (N=14,381).

|  | Exposed to any<br>BZRA<br>(N=686) | Not exposed to<br>any BZRA<br>(N=13,695) | Non-exposed<br>matched group<br>(N=686) | Exposed to any BZRA<br>vs.<br>Not exposed | Exposed to any BZRA<br>vs.<br>Not exposed | Exposed to any<br>BZRA<br>vs.<br>Non-exposed<br>matched group |
| --- | --- | --- | --- | --- | --- | --- |
|  |  |  |  | Crude analysis | Analysis weighted by<br>inverse-probability-weighting weights | Matched analytic<br>sample analysis |
|  | N (%) | N (%) | N (%) | SMD | SMD | SMD |
| Age |  |  |  | 0.893 | 0.174 | 0.029 |
| 18 to 50 years | 70 (10.2%) | 5669 (41.4%) | 65 (9.4%) |  |  |  |
| 51 to 70 years | 192 (28.0%) | 4395 (32.1%) | 198 (28.9%) |  |  |  |
| More than 70 years | 424 (61.8%) | 3631 (26.5%) | 423 (61.7%) |  |  |  |
| Sex |  |  |  | 0.047 | 0.095 | 0.108 |
| Women | 348 (50.7%) | 7270 (53.1%) | 311 (45.3%) |  |  |  |
| Men | 338 (49.3%) | 6425 (46.9%) | 375 (54.7%) |  |  |  |
| Hospital |  |  |  | 0.387 | 0.050 | 0.063 |
| *AP-HP Centre – Paris University,*<br>*Henri Mondor University Hospitals and*<br>*at home hospitalization* | 206 (30.0%) | 6585 (48.1%) | 212 (30.9%) |  |  |  |
| *AP-HP Nord and Hôpitaux*<br>*Universitaires Paris Seine-Saint-Denis* | 222 (32.4%) | 3685 (26.9%) | 227 (33.1%) |  |  |  |
| *AP-HP Paris Saclay University* | 122 (17.8%) | 1575 (11.5%) | 106 (15.5%) |  |  |  |
| *AP-HP Sorbonne University* | 136 (19.8%) | 1850 (13.5%) | 141 (20.6%) |  |  |  |
| Obesity <sup>a</sup> |  |  |  | 0.186 | 0.074 | 0.079 |
| Yes | 135 (19.7%) | 1758 (12.8%) | 114 (16.6%) |  |  |  |
| No | 551 (80.3%) | 11937 (87.2%) | 572 (83.4%) |  |  |  |
| Smoking <sup>b</sup> |  |  |  | 0.263 | 0.074 | 0.065 |
| Yes | 112 (16.3%) | 1072 (7.83%) | 96 (14.0%) |  |  |  |
| No | 574 (83.7%) | 12623 (92.2%) | 590 (86.0%) |  |  |  |
| Any medical condition <sup>c</sup> |  |  |  | 0.711 | 0.058 | 0.027 |
| Yes | 393 (57.3%) | 3336 (24.4%) | 402 (58.6%) |  |  |  |
| No | 293 (42.7%) | 10359 (75.6%) | 284 (41.4%) |  |  |  |
| Medication according to compassionate<br>use or as part of a clinical trial <sup>d</sup> |  |  |  | 0.371 | 0.103 | 0.017 |
| Yes | 170 (24.8%) | 1483 (10.8%) | 165 (24.1%) |  |  |  |
| No | 516 (75.2%) | 12212 (89.2%) | 521 (75.9%) |  |  |  |
| Any current psychiatric disorder <sup>e</sup> |  |  |  | 0.582 | 0.052 | 0.007 |
| Yes | 154 (22.4%) | 498 (3.6%) | 152 (22.2%) |  |  |  |
| *No* | 532 (77.6%) | 13197 (96.4%) | 534 (77.8%) |  |  |  |
| Any antidepressant |  |  |  | 1.045 | 0.185 | 0.075 |
| *Yes* | 285 (41.5%) | 411 (3.0%) | 260 (37.9%) |  |  |  |
| *No* | 401 (58.5%) | 13284 (97.0%) | 426 (62.1%) |  |  |  |
| Any mood stabilizer medication <sup>f</sup> |  |  |  | 0.617 | 0.404 | 0.011 |
| *Yes* | 143 (20.8%) | 284 (2.1%) | 140 (20.4%) |  |  |  |
| *No* | 543 (79.2%) | 13411 (97.9%) | 546 (79.6%) |  |  |  |
| Any antipsychotic medication |  |  |  | 0.714 | 0.305 | 0.014 |
| *Yes* | 163 (23.8%) | 198 (1.4%) | 159 (23.2%) |  |  |  |
| *No* | 523 (76.2%) | 13497 (98.6%) | 527 (76.8%) |  |  |  |

<sup>a</sup> Defined as having a body-mass index higher than 30 kg/m<sup>2</sup> or an International Statistical Classification of Diseases and Related Health Problems (ICD-10) diagnosis code for obesity (E66.0, E66.1, E66.2, E66.8, E66.9).
<sup>b</sup> Current Smoking status was self-reported.
<sup>c</sup> Assessed using ICD-10 diagnosis codes for diabetes mellitus (E11), diseases of the circulatory system (I00-I99), diseases of the respiratory system (J00-J99), neoplasms (C00-D49), diseases of the blood and blood-forming organs and certain disorders involving the immune mechanism (D5-D8), frontotemporal dementia (G31.0), peptic ulcer (K27), diseases of liver (K70-K95), hemiplegia or paraplegia (G81-G82), acute kidney failure or chronic kidney disease (N17-N19), and HIV (B20).
<sup>d</sup> Any medication prescribed as part of a clinical trial or according to compassionate use (e.g., hydroxychloroquine, azithromycin, remdesivir, tocilizumab, sarilumab or dexamethasone).
<sup>e</sup> Assessed using ICD-10 diagnosis codes (F00- F99).
<sup>f</sup> Included lithium and antiepileptic medications with mood stabilizing properties.
SMD>0.1 indicates substantial difference.
Abbreviation: SMD, standardized mean difference.

### 3.2. Study endpoint

The endpoint of death occurred in 186 patients (27.1%) who received a BZRA at baseline and 1,134 patients (8.3%) who did not. The crude, unadjusted analysis (hazard ratio (HR=3.20; 95% CI=2.74-3.74; p<0.001), the primary analysis with inverse probability weighting (HR=1.61; 95% CI=1.31-1.98; p<0.001) and the multivariable IPW Cox regression adjusting for unbalanced covariates (HR=1.56; 95% CI=1.29-1.89; p<0.001) showed a significant association between BZRA use and increased mortality (**Figure 1**; **Table 2**). A post-hoc analysis indicated that we had 80% power in the crude analysis to detect a hazard ratio of at least 1.27.

**Figure 1.**
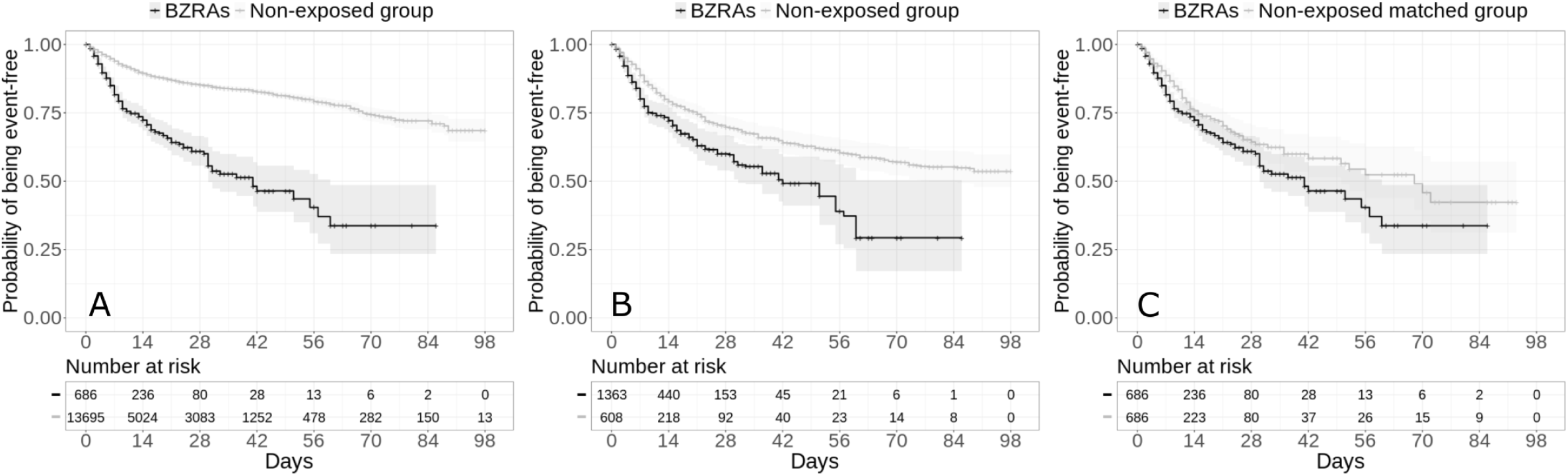
Kaplan-Meier curves for mortality in the full sample crude analysis (N=14,381) (A), in the full sample analysis with IPW (N=14,381) (B), and in the matched analytic sample (N=1,372) (C), according to BZRA use at baseline. The shaded areas represent pointwise 95% confidence intervals.

**Table 2.**
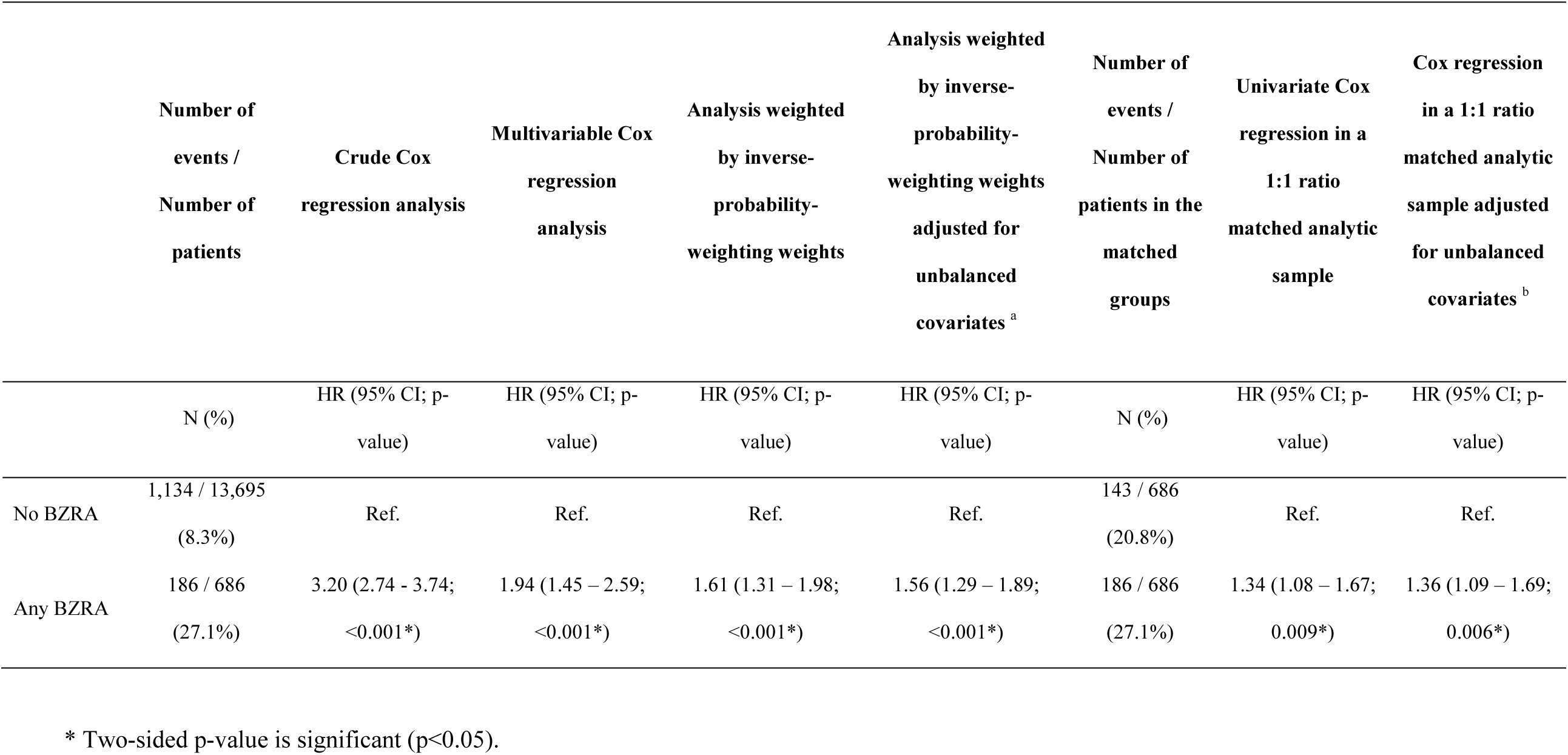
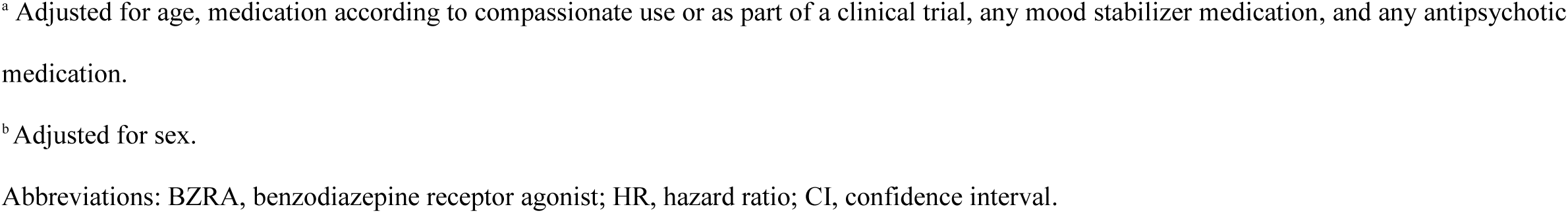
Association between benzodiazepine receptor agonist (BZRA) use at baseline and the endpoint of death.

|  | Number of<br>events /<br>Number of<br>patients | Crude Cox<br>regression analysis | Multivariable Cox<br>regression<br>analysis | Analysis weighted<br>by inverse-probability-weighting weights | Analysis weighted<br>by inverse-probability-weighting weights<br>adjusted for<br>unbalanced<br>covariates <sup>a</sup> | Number of<br>events /<br>Number of<br>patients in the<br>matched<br>groups | Univariate Cox<br>regression in a<br>1:1 ratio<br>matched analytic<br>sample | Cox regression<br>in a 1:1 ratio<br>matched analytic<br>sample adjusted<br>for unbalanced<br>covariates <sup>b</sup> |
| --- | --- | --- | --- | --- | --- | --- | --- | --- |
|  | N (%) | HR (95% CI; p-value) | HR (95% CI; p-value) | HR (95% CI; p-value) | HR (95% CI; p-value) | N (%) | HR (95% CI; p-value) | HR (95% CI; p-value) |
| No BZRA | 1,134 / 13,695<br>(8.3%) | Ref. | Ref. | Ref. | Ref. | 143 / 686<br>(20.8%) | Ref. | Ref. |
| Any BZRA | 186 / 686<br>(27.1%) | 3.20 (2.74 - 3.74;<br><0.001*) | 1.94 (1.45 – 2.59;<br><0.001*) | 1.61 (1.31 – 1.98;<br><0.001*) | 1.56 (1.29 – 1.89;<br><0.001*) | 186 / 686<br>(27.1%) | 1.34 (1.08 – 1.67;<br>0.009*) | 1.36 (1.09 – 1.69;<br>0.006*) |

\* Two-sided p-value is significant (p<0.05).
<sup>a</sup> Adjusted for age, medication according to compassionate use or as part of a clinical trial, any mood stabilizer medication, and any antipsychotic medication.
<sup>b</sup> Adjusted for sex.
Abbreviations: BZRA, benzodiazepine receptor agonist; HR, hazard ratio; CI, confidence interval.

In sensitivity analyses, the multivariable Cox regression model in the full sample also a significant association (HR=1.94; 95% CI=1.45-2.59; p<0.001), as did the univariate Cox regression model in a matched analytic sample using a 1:1 ratio (HR=1.34; 95% CI=1.08-1.67; p=0.009) (**Table 2**). Similarly, the primary analysis using imputed data yielded significant results (**eTable 2**), as did that considering BZRA use as a time-dependent variable and including all patients who received a BZRA at any time during the visit (**eTable 3**). The exclusion from the analyses of the patients who had been admitted to ICUs did not alter the significance of the association (**eTable 4**).

Additional analyses showed a significant dose-dependent relationship between baseline daily BZRA dose and the endpoint (HR=1.55; 95% CI=1.08-2.22; p=0.017), based on the primary IPW analysis. Additional adjustments for respiratory depression, any other clinical markers of disease severity, or both, resulted in still significant associations, which were of similar magnitude to that observed in the primary analysis (**eTable 5**). We found that all individual BZRAs were significantly associated with increased mortality, except for diazepam (**eTable 6**). Following adjustments, there were significant associations of any BZRA other than diazepam, midazolam, and any BZRA other than diazepam and midazolam with increased mortality, as compared to not receiving BZRAs (**eTable 6**). Finally, compared with any other BZRA treatment, diazepam use was significantly associated with reduced mortality in the crude analysis (HR, 0.44; 95% CI, 0.23 to 0.86; p=0.017), in the primary analysis (HR, 0.31; 95% CI, 0.13 to 0.74; p=0.008), and in the sensitivity analyses (**eFigure 2; eTables 6 to8**).

## 4. Discussion

In this multicenter retrospective observational study involving a large sample of patients hospitalized for COVID-19, we found that benzodiazepine receptor agonist use were significantly and substantially associated with increased mortality, independently of patient characteristics and other medications, and with a significant dose-dependent relationship. This association remained significant in multiple sensitivity analyses. Secondary exploratory analyses suggested that most BZRAs could be associated with this risk in these patients.

We found that BZRA use was significantly associated with increased mortality among patients hospitalized for COVID-19. This association might be explained by several mechanisms. First, although the risk of respiratory depression associated with benzodiazepines in the general population is debated and was not found to play a substantial role in our study, it might be relevant among elderly patients with COVID-19 and in those with pre-existing comorbidities, such as chronic obstructive pulmonary disease (35). Second, benzodiazepines may be associated with an increased risk of secondary infections, and particularly pneumonia, in patients with COVID-19 (36). Finally, in patients with COVID-19 and known risk factors for delirium (e.g., old age, dementia, multiple comorbidities), BZRA use may favor the occurrence of this condition (35), which is associated with unfavorable COVID-19 disease prognosis (37). These results suggest the need to carefully reevaluate the indication of BZRAs and decrease in dose or taper these medications when possible in patients with COVID-19, in line with professional guidelines recommendation for most patients on long-term benzodiazepines (4).

Secondary exploratory analyses indicate that most BZRAs may be associated with increased mortality among patients hospitalized for COVID-19, except diazepam. Interestingly, this result could be in line with preclinical findings. A prior study indicated a role of acid sphingomyelinase/ceramide system in SARS-CoV-2 infections (38) and that the formation of ceramide-enriched membrane platforms may mediate the entry of the virus into epithelial cells. Although most BZRAs do not influence cellular ceramide levels, diazepam has been shown to interact with the acyl coenzyme A binding protein, also known as ‘diazepam binding inhibitor’, a protein that contributes to ceramide synthesis (39,40). This interaction may result in a reduced activity of the ceramide synthase pathway and, finally, reduced cellular ceramide concentration, which might possibly explain reduced adverse effects of diazepam on COVID-19 compared to other BZRAs. However, given the exploratory nature of these analyses, the observational design of the study and the long half-life of diazepam, this result should be interpreted with caution and other studies are required to confirm this finding and this potential underlying mechanism.

Our study has several limitations. First, there are two possible major inherent biases in observational studies: unmeasured confounding and confounding by indication. We tried to minimize the effects of confounding in several different ways. First, we used an analysis with inverse probability weighting to minimize the effects of confounding by indication (19,28). Second, we performed multiple sensitivity analyses, which showed similar results. Finally, although some amount of unmeasured confounding may remain, our analyses adjusted for numerous potential confounders. Other limitations include missing data for some baseline characteristic variables (i.e., 11.5%), which might be explained by the overwhelming of all hospital units during the COVID-19 peak incidence, and different results might have been observed during a lower COVID-19 incidence period. However, imputation of missing data did not alter the significance of our results. Second, inflation of type I error might have occurred in secondary analyses due to multiple testing. Third, our study cannot establish a causal relationship between BZRA use and increased mortality. Finally, despite the multicenter design, our results may not be generalizable to outpatients or other regions.

In this multicenter observational retrospective study, benzodiazepine receptor agonist use was significantly and substantially associated with increased mortality among adult patients hospitalized for COVID-19, with a significant dose-dependent relationship. This finding suggests a potential benefit of decreasing dose or tapering off these medications when possible in these patients. Other studies are needed to confirm this result and determine the exact mechanisms underlying this association.

## Data Availability

Data from the AP-HP Health Data Warehouse can be obtained with permission at https://eds.aphp.fr//.

## ACKNOWLEDGMENT SECTION

### Contributions

NH designed the study, performed statistical analyses, and wrote the first draft of the manuscript. MSR contributed to study design, performed statistical analyses and critically revised the manuscript. FL contributed to study design and critically revised the manuscript for scientific content. RV contributed to statistical analyses and critically revised the manuscript for scientific content. All other authors critically revised the manuscript for scientific content.

### Data Availability Statement

Data from the AP-HP Health Data Warehouse can be obtained with permission at https://eds.aphp.fr//.

### Funding source

This work did not receive any external funding.

### Disclaimer

The information contained in this study is provided for research purpose and should not be used as a substitute or replacement for diagnosis or treatment recommendations or other clinical decisions or judgment. The views presented in this manuscript are those of the authors and do not necessarily represents those of the National Institute on Drug Abuse or any U.S. Government Agency.

## Acknowledgment

The authors thank the EDS APHP Covid consortium integrating the APHP Health Data Warehouse team as well as all the APHP staff and volunteers who contributed to the implementation of the EDS-Covid database and operating solutions for this database. Collaborators of the EDS APHP Covid consortium: Pierre-Yves ANCEL, Alain BAUCHET, Nathanaël BEEKER, Vincent BENOIT, Mélodie BERNAUX, Ali BELLAMINE, Romain BEY, Aurélie BOURMAUD, Stéphane BREANT, Anita BURGUN, Fabrice CARRAT, Charlotte CAUCHETEUX, Julien CHAMP, Sylvie CORMONT, Christel DANIEL, Julien DUBIEL, Catherine DUCLOAS, Loic ESTEVE, Marie FRANK, Nicolas GARCELON, Alexandre GRAMFORT, Nicolas GRIFFON, Olivier GRISEL, Martin GUILBAUD, Claire HASSEN-KHODJA, François HEMERY, Martin HILKA, Anne Sophie JANNOT, Jerome LAMBERT, Richard LAYESE, Judith LEBLANC, Léo LEBOUTER, Guillaume LEMAITRE, Damien LEPROVOST, Ivan LERNER, Kankoe LEVI SALLAH, Aurélien MAIRE, Marie-France MAMZER, Patricia MARTEL, Arthur MENSCH, Thomas MOREAU, Antoine NEURAZ, Nina ORLOVA, Nicolas PARIS, Bastien RANCE, Hélène RAVERA, Antoine ROZES, Elisa SALAMANCA, Arnaud SANDRIN, Patricia SERRE, Xavier TANNIER, Jean-Marc TRELUYER, Damien VAN GYSEL, Gaël VAROQUAUX, Jill Jen VIE, Maxime WACK, Perceval WAJSBURT, Demian WASSERMANN, Eric ZAPLETAL.

