## Supplementary material for "Association between benzodiazepine receptor agonist use and increased mortality among patients hospitalized for COVID-19: results from an observational study"

### SUPLEMENTARY MATERIAL


eMethods 2

eFigure 1. Study cohort. 3

eFigure 2. Kaplan-Meier curves for mortality in the full sample crude analysis (N=686) (A), in the full sample analysis with inverse probability weighting (N=686) (B) and in the matched analytic sample using a 1:1 ratio (N=148) (C) of patients who had been hospitalized for COVID-19, according to diazepam or other benzodiazepine receptor agonist (BZRA) use at baseline. 4

eTable 1. Associations of baseline characteristics with the endpoint of death in the cohort of adult patients who had been hospitalized for COVID-19 (N=14,381). 5

eTable 2. Association between benzodiazepine receptor agonist (BZRA) use at baseline and mortality in models imputing missing data using multiple imputation. 8

eTable 3. Associations between BZRA use and mortality in patients hospitalized for COVID-19, when including all patients who received a BZRA and considering BZRA use as a time-dependent variable. 9

eTable 4. Association between benzodiazepine receptor agonist (BZRA) use and the endpoint of death among patients who had been hospitalized for COVID-19 outside ICUs (N = 13,693). 10

eTable 5. Association between benzodiazepine receptor agonist (BZRA) use at baesline and mortality, following additional adjustments for respiratory depression, any other clinical markers of disease severity, or both. 11

eTable 6. Associations between individual benzodiazepine receptor agonists (BZRAs) and mortality. 13

eTable 7. Characteristics of patients with COVID-19 receiving diazepam versus any other benzodiazepine receptor agonist (BZRA) (N=686). 14

eTable 8. Association between diazepam use and the endpoint of death among patients who had been hospitalized for COVID-19 and had received benzodiazepine receptor agonists at baseline (N=686). 16

**eMethods**

We obtained the following data for each patient at the time of the hospitalization: sex; age, which was categorized into 3 classes based on the OpenSAFELY study results use[22] (i.e. 18-50, 51-70, 71+); hospital, which was categorized into 4 classes following the administrative clustering of AP-HP hospitals in Paris and its suburbs based on their geographical location (i.e., AP-HP Centre – Paris University, Henri Mondor University Hospitals and at home hospitalization; AP-HP Nord and Hôpitaux Universitaires Paris Seine-Saint-Denis; AP-HP Paris Saclay University; and AP-HP Sorbonne University); obesity, which was defined as having a body mass index higher than 30 kg/m$^2$ or an International Statistical Classification of Diseases and Related Health Problems (ICD-10) diagnosis code for obesity (E66.0, E66.1, E66.2, E66.8, E66.9); self-reported current smoking status; any medical condition associated with increased clinical severity related to COVID-19 or benzodiazepine receptor agonist use [22–25], based on ICD-10 diagnosis codes, including diabetes mellitus (E11), diseases of the circulatory system (I00-I99), diseases of the respiratory system (J00-J99), neoplasms (C00-D49), diseases of the blood and blood-forming organs and certain disorders involving the immune mechanism (D5-D8), frontotemporal dementia (G31.0), peptic ulcer (K27), diseases of liver (K70-K95), hemiplegia or paraplegia (G81-G82), acute kidney failure or chronic kidney disease (N17-N19) and HIV (B20); any medication prescribed according to compassionate use or as part of a clinical trial (e.g. hydroxychloroquine, azithromycin, remdesivir, tocilizumab, sarilumab, or dexamethasone); and clinical markers of disease severity, including respiratory depression, defined by a respiratory rate < 12 breaths/min or a resting peripheral capillary oxygen saturation in ambient air < 90%[27], and any other clinical marker of disease severity, defined as having temperature > 40°C or systolic blood pressure < 100 mmHg or respiratory rate > 24 breaths/min or plasma lactate levels higher than 2 mmol/L[27]. To take into account possible confounding by indication bias for BZRAs, we recorded whether patients had any current psychiatric disorder (F00-F99) based on ICD-10 diagnosis codes, and whether they were prescribed any other psychotropic medication, including any antidepressant, mood stabilizer (i.e. lithium or antiepileptic medications with mood stabilizing effects), or antipsychotic medication.

All medical notes and prescriptions are computerized in Greater Paris University hospitals. Medications including their dosage, frequency, date, and mode of administration were identified from medication administration data or scanned hand-written medical prescriptions, through two deep learning models based on BERT contextual embeddings [18], one for the medications and another for their mode of administration. The model was trained on the APmed corpus [19], a previously annotated dataset for this task. Extracted medications names were then normalized to the Anatomical Therapeutic Chemical (ATC) terminology using approximate string matching.

**eFigure 1. Study cohort.**

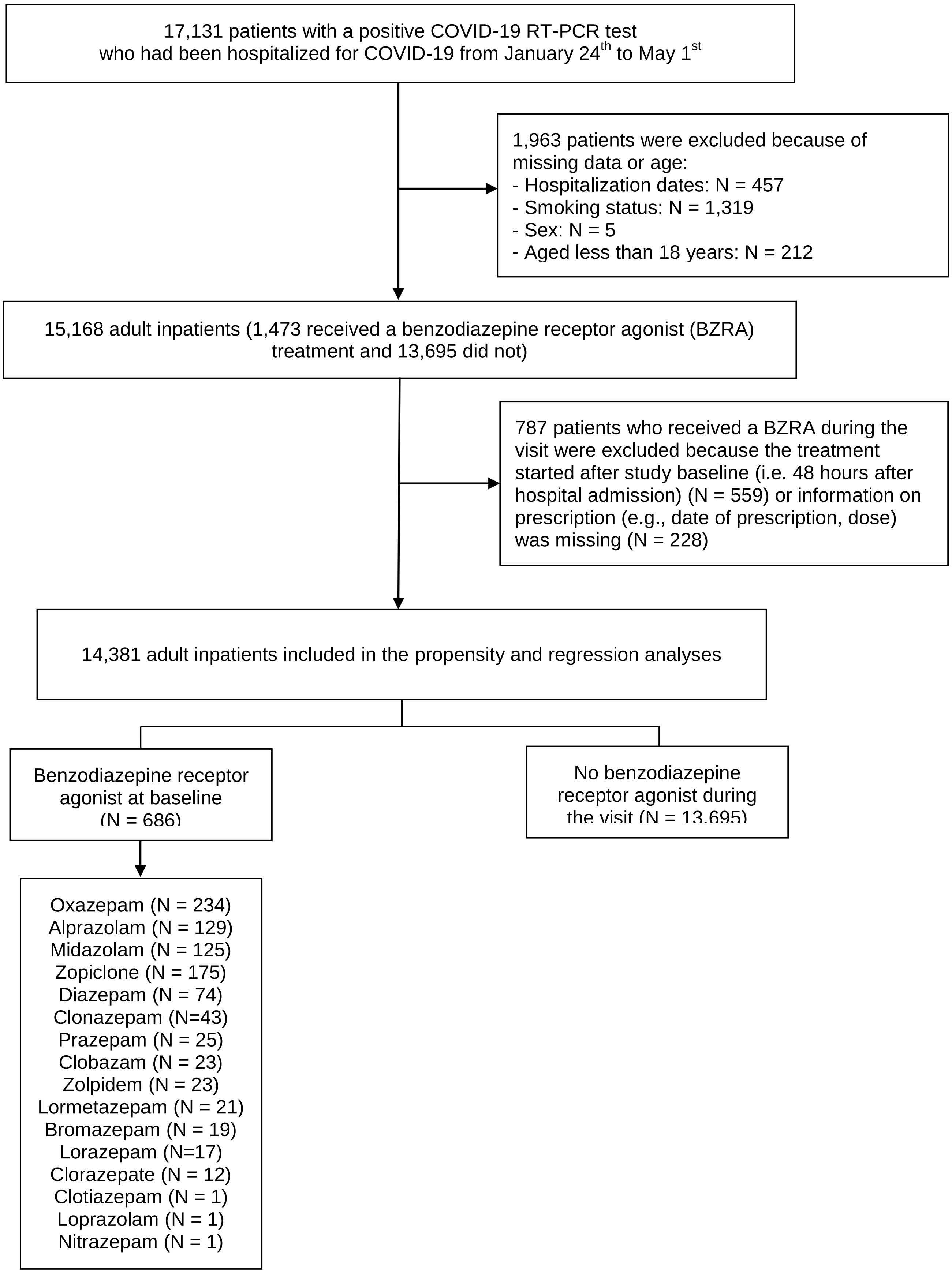

**eFigure 2. Kaplan-Meier curves for mortality in the full sample crude analysis (N=686) (A), in the full sample analysis with inverse probability weighting (N=686) (B) and in the matched analytic sample using a 1:1 ratio (N=148) (C) of patients who had been hospitalized for COVID-19, according to diazepam or other benzodiazepine receptor agonist (BZRA) use at baseline.**

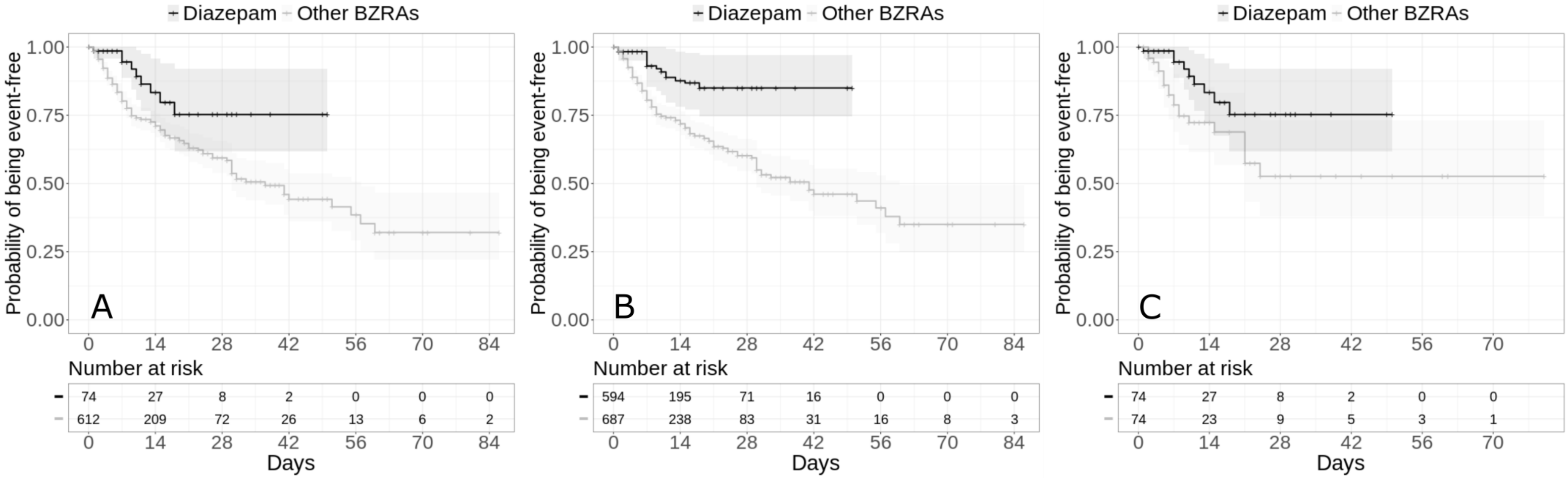


The shaded areas represent pointwise 95% confidence intervals.

**eTable 1. Associations of baseline characteristics with the endpoint of death in the cohort of adult patients who had been hospitalized for COVID-19 (N=14,381).**

| | | | | Endpoint of death | | |
|---|---|---|---|---|---|---|
| | **Full population (N =14,381)** | **With the end-point event (N =1,320)** | **Without the end-point event (N =13,061)** | **Crude analysis** | **Multivariable analysis µ** | |
| | **N (%)** | **N (%)** | **N (%)** | **HR (95% CI; p-value)** | **HR (95% CI; p-value)** | **Collinearity diagnostics (variance inflation factor)** |
| Age | | | | | | 1.02 |
| *18 to 50 years* | 5739 (39.9%) | 47 (3.56%) | 5692 (43.6%) | Ref. | Ref. | |
| *51 to 70 years* | 4587 (31.9%) | 322 (24.4%) | 4265 (32.7%) | 8.23 (6.06 - 11.18; <0.001*) | 3,14 (1,46 - 6,76; 0,003*) | |
| *More than 70 years* | 4055 (28.2%) | 951 (72.0%) | 3104 (23.8%) | 24.79 (18.49 - 33.25; <0.001*) | 7,44 (3,70 - 14,97; <0,001*) | |
| Sex | | | | | | 1.05 |
| *Women* | 7618 (53.0%) | 484 (36.7%) | 7134 (54.6%) | Ref. | Ref. | |
| *Men* | 6763 (47.0%) | 836 (63.3%) | 5927 (45.4%) | 2.02 (1.81 - 2.26; <0.001*) | 1,72 (1,31 - 2,24; <0,001*) | |
| Hospital | | | | | | 1.03 |
| *AP-HP Centre – Paris University, Henri Mondor University Hospitals and at home hospitalization* | 6791 (47.2%) | 412 (31.2%) | 6379 (48.8%) | Ref. | Ref. | |
| *AP-HP Nord and Hôpitaux Universitaires Paris Seine-Saint-Denis* | 3907 (27.2%) | 450 (34.1%) | 3457 (26.5%) | 2.28 (1.99 - 2.6; <0.001*) | 1,69 (1,25 - 2,28; 0,001*) | |

| | | | | | | |
|---|---|---|---|---|---|---|
| *AP-HP Paris Saclay University* | 1697 (11.8%) | 231 (17.5%) | 1466 (11.2%) | 2.24 (1.91 - 2.64; <0.001*) | 1,06 (0,70 - 1,63; 0,771) | |
| *AP-HP Sorbonne University* | 1986 (13.8%) | 227 (17.2%) | 1759 (13.5%) | 2.01 (1.71 - 2.37; <0.001*) | 0,96 (0,70 - 1,33; 0,821) | |
| Obesity [α] | | | | | | 1.03 |
| *Yes* | 1893 (13.2%) | 279 (21.1%) | 1614 (12.4%) | 1.54 (1.35 - 1.76; <0.001*) | 1,04 (0,77 - 1,40 - 0,818) | |
| *No* | 12488 (86.8%) | 1041 (78.9%) | 11447 (87.6%) | Ref. | Ref. | |
| Smoking [β] | | | | | | 1.02 |
| *Yes* | 1184 (8.2%) | 199 (15.1%) | 985 (7.54%) | 1.73 (1.49 - 2.01; <0.001*) | 0,80 (0,54 - 1,18; 0,259) | |
| *No* | 13197 (91.8%) | 1121 (84.9%) | 12076 (92.5%) | Ref. | Ref. | |
| Any medical condition [γ] | | | | | | 1.20 |
| *Yes* | 3729 (25.9%) | 903 (68.4%) | 2826 (21.6%) | 7.25 (6.43 - 8.16; <0.001*) | 3,17 (2,36 - 4,27; <0,001*) | |
| *No* | 10652 (74.1%) | 417 (31.6%) | 10235 (78.4%) | Ref. | Ref. | |
| Medication according to compassionate use or as part of a clinical trial [ϴ] | | | | | | 1.04 |
| *Yes* | 1653 (11.5%) | 277 (21.0%) | 1376 (10.5%) | 1.94 (1.7 - 2.22; <0.001*) | 0,92 (0,74 - 1,15; 0,476) | |
| *No* | 12728 (88.5%) | 1043 (79.0%) | 11685 (89.5%) | Ref. | Ref. | |
| Any current psychiatric disorder [¥] | | | | | | 1.13 |
| *Yes* | 652 (4.5%) | 240 (18.2%) | 412 (3.15%) | 4.86 (4.22 - 5.6; <0.001*) | 1,57 (1,20 - 2,06; 0,001*) | |
| *No* | 13729 (95.5%) | 1080 (81.8%) | 12649 (96.8%) | Ref. | Ref. | |
| Any antidepressant | | | | | | 1.07 |
| *Yes* | 696 (4.8%) | 155 (11.7%) | 541 (4.1%) | 2.27 (1.92 - 2.69; <0.001*) | 1,05 (0,76 - 1,44; 0,783) | |
| *No* | 13685 (95.2%) | 1165 (88.3%) | 12520 (95.9%) | Ref. | Ref. | |
| Any mood stabilizer medication [Ω] | | | | | | 1.03 |
| *Yes* | 427 (2.97%) | 77 (5.8%) | 350 (2.7%) | 1.74 (1.38 - 2.19; <0.001) | 0,91 (0,58 - 1,41; 0,660) | |

| | | | | | | |
|---|---|---|---|---|---|---|
| *No* | 13954 (97.0%) | 1243 (94.2%) | 12711 (97.3%) | Ref. | Ref. | |
| Any antipsychotic medication | | | | | | 1.04 |
| *Yes* | 361 (2.5%) | 65 (4.9%) | 296 (2.3%) | 1.77 (1.38 - 2.27; <0.001*) | 0,73 (0,44 - 1,21; 0,225) | |
| *No* | 14020 (97.5%) | 1255 (95.1%) | 12765 (97.7%) | Ref. | Ref. | |
| Respiratory depression [£] | | | | | | 1.45 |
| *Yes* | 387 (2.69%) | 153 (11.6%) | 234 (1.79%) | 2.5 (2.09 - 2.99; <0.001*) | 1,49 (1,07 - 2,06; 0,017*) | |
| *No* | 3560 (24.8%) | 547 (41.4%) | 3013 (23.1%) | Ref. | Ref. | |
| *Missing* | 10434 (72.6%) | 620 (47.0%) | 9814 (75.1%) | 0.32 (0.28 - 0.36; <0.001*) | 1,16 (0,87 - 1,55; 0,311) | |
| Any other clinical marker of severity [§] | | | | | | 1.45 |
| *Yes* | 2079 (14.5%) | 526 (39.8%) | 1553 (11.9%) | 2.13 (1.84 - 2.47; <0.001*) | 2,08 (1,59 - 2,71; <0,001*) | |
| *No* | 2282 (15.9%) | 263 (19.9%) | 2019 (15.5%) | Ref. | Ref. | |
| *Missing* | 10020 (69.7%) | 531 (40.2%) | 9489 (72.7%) | 0.38 (0.33 - 0.44; <0.001*) | 0,83 (0,59 - 1,17; 0,284) | |

[α] Defined as having a body-mass index higher than 30 $kg/m^2$ or an International Statistical Classification of Diseases and Related Health Problems (ICD-10) diagnosis code for obesity (E66.0, E66.1, E66.2, E66.8, E66.9).
[β] Current Smoking status was self-reported.
[γ] Assessed using ICD-10 diagnosis codes for diabetes mellitus (E11), diseases of the circulatory system (I00-I99), diseases of the respiratory system (J00-J99), neoplasms (C00-D49), diseases of the blood and blood-forming organs and certain disorders involving the immune mechanism (D5-D8), frontotemporal dementia (G31.0), peptic ulcer (K27), diseases of liver (K70-K95), hemiplegia or paraplegia (G81-G82), acute kidney failure or chronic kidney disease (N17-N19), and HIV (B20).
[ϴ] Any medication prescribed as part of a clinical trial or according to compassionate use (e.g., hydroxychloroquine, azithromycin, remdesivir, tocilizumab, sarilumab or dexamethasone).
[¥] Assessed using ICD-10 diagnosis codes (F00-F99).
[Ω] Included lithium and antiepileptic medications with mood stabilizing properties.
[µ] Adjusted for sex, age, hospital type, obesity, current smoking status, any significant medical or psychiatric condition, any medication prescribed according to compassionate use or as part of a clinical trial and other psychotropic medications (i.e. any antidepressant, mood stabilizer and antipsychotic medication).
[£] Defined by a respiratory rate < 12 breaths/min or a resting peripheral capillary oxygen saturation in ambient air < 90%,
[§] Defined by a temperature > 40°C or a systolic blood pressure < 100 mmHg or a respiratory rate > 24 breaths/min or a plasma lactate levels higher than 2 mmol/L
* two-sided p-value is significant (p<0.05).
Abbreviations: HR, hazard ratio; SE, standard error; VIF, variance inflation factor.

**eTable 2. Association between benzodiazepine receptor agonist (BZRA) use at baseline and mortality in models imputing missing data using multiple imputation.**

| | Number of events / Number of patients | Crude Cox regression analysis | Multivariable Cox regression analysis | Analysis weighted by inverse-probability-weighting weights | Analysis weighted by inverse-probability-weighting weights adjusted for unbalanced covariates [a] | Number of events / Number of patients in the matched groups | Univariate Cox regression in a 1:1 ratio matched analytic sample |
|---|---|---|---|---|---|---|---|
| | N (%) | HR (95% CI; p-value) | HR (95% CI; p-value) | HR (95% CI; p-value) | HR (95% CI; p-value) | N (%) | HR (95% CI; p-value) |
| No BZRA | 1,299 / 15,602 (8.3%) | Ref. | Ref. | Ref. | Ref. | 148 / 705 (21.0%) | Ref. |
| Any BZRA | 190 / 705 (27.0%) | 3.15 (2.71 - 3.67; <0.001*) | 1.82 (1.33 – 2.48; <0.001*) | 1.51 (1.23 – 1.84; <0.001*) | 1.52 (1.28 – 1.81; <0.001*) | 190 / 705 (27.0%) | 1.30 (1.05 – 1.62; 0.016*) |

* Two-sided p-value is significant (p<0.05).
[a] Adjusted for age, any medical condition, any mood stabilizer medication, current psychiatric disorder, any antidepressant, any mood stabilizer medication, and any antipsychotic medication.
Abbreviations: HR, hazard ratio; CI, confidence interval.

**eTable 3. Associations between BZRA use and mortality in patients hospitalized for COVID-19, when including all patients who received a BZRA and considering BZRA use as a time-dependent variable.**

| | Number of events / Number of patients | Crude Cox regression analysis | Multivariable Cox regression analysis | Analysis weighted by inverse-probability-weighting weights | Analysis weighted by inverse-probability-weighting weights adjusted for unbalanced covariates [a] | Number of events / Number of patients in the matched groups | Crude Cox regression analysis |
|---|---|---|---|---|---|---|---|
| | N (%) | HR (95% CI; p-value) | HR (95% CI; p-value) | HR (95% CI; p-value) | HR (95% CI; p-value) | N (%) | HR (95%CI; p-value) |
| No BZRA | 1,134 / 13,695 (8.3%) | Ref. | Ref. | Ref. | Ref. | 230 / 1,089 (21.1%) | Ref. |
| Any BZRA | 248 / 1,089 (22.8%) | 4.41 (4.11 – 5.41; <0.001*) | 2.59 (2.22 – 3.01; <0.001*) | 1.84 (1.53 – 2.23; <0.001*) | 1.55 (1.26 – 1.90; <0.001*) | 248 / 1,089 (22.8%) | 1.18 (0.87 – 1.60; 0.290) |

* Two-sided p-value is significant (p<0.05).
[a] Adjusted for age, sex, hospital, medication according to compassionate use or as part of a clinical trial, any mood stabilizer medication, and any antipsychotic medication, any current psychiatric disorder and any mood stabilizer medication.
Abbreviations: HR, hazard ratio; CI, confidence interval.

**eTable 4. Association between benzodiazepine receptor agonist (BZRA) use and the endpoint of death among patients who had been hospitalized for COVID-19 outside ICUs (N = 13,693).**

| | Number of events / Number of patients | Crude Cox regression analysis | Multivariable Cox regression analysis | Analysis weighted by inverse-probability-weighting weights | Analysis weighted by inverse-probability-weighting weights adjusted for unbalanced covariates | Number of events / Number of patients in the matched groups | Crude Cox regression analysis |
|---|---|---|---|---|---|---|---|
| | N (%) | HR (95%CI; p-value) | HR (95%CI; p-value) $^{\alpha}$ | HR (95%CI; p-value) $^{\alpha}$ | HR (95%CI; p-value) $^{\alpha}$ | N (%) | HR (95%CI; p-value) |
| No BZRA | 900 / 13,068 (6.9%) | Ref. | Ref. | Ref. | Ref. | 127 / 625 (20.3%) | Ref. |
| Any BZRA | 165 / 625 (26.4%) | 3.62 (2.72 – 4.80; <0.001*) | 2.10 (1.50 – 2.95; <0.001*) | 1.73 (1.39 – 2.15; <0.001*) | 1.73 (1.39 – 2.15; <0.001*) | 165 / 625 (26.4%) | 1.34 (1.06 – 1.69; 0.013*) |

* Two-sided p-value is significant (p<0.05).
Abbreviations: HR, hazard ratio; CI, confidence interval.

**eTable 5. Association between benzodiazepine receptor agonist (BZRA) use at baesline and mortality, following additional adjustments for respiratory depression, any other clinical markers of disease severity, or both.**

| | **Number of events / Number of patients** | **Multivariable Cox regression analysis** | **Analysis weighted by inverse-probability-weighting weights** | **Analysis weighted by inverse-probability-weighting weights adjusted for unbalanced covariates** | **Number of events / Number of patients in the matched groups** | **Univariate Cox regression in a 1:1 ratio matched analytic sample** |
|---|---|---|---|---|---|---|
| | N (%) | HR (95% CI; p-value) | HR (95% CI; p-value) | HR (95% CI; p-value) | N (%) | HR (95% CI; p-value) |
| Main analyses without adjustments for respiratory depression or any other clinical markers of disease severity | | | | | | |
| No BZRA | 1,134 / 13,695 (8.3%) | Ref. | Ref. | Ref. | 143 / 686 (20.8%) | Ref. |
| Any BZRA | 186 / 686 (27.1%) | 1.94 (1.45 – 2.59; <0.001*) | 1.61 (1.31 – 1.98; <0.001*) | 1.56 (1.29 – 1.89; <0.001*) | 186 / 686 (27.1%) | 1.34 (1.08 – 1.67; 0.009*) |
| Analyses adjusting in addition for respiratory depression | | | | | | |
| No BZRA | 1,134 / 13,695 (8.3%) | Ref. | Ref. | Ref. | 144 / 686 (21.0%) | Ref. |
| Any BZRA | 186 / 686 (27.1%) | 1.86 (1.34 – 2.50; <0.001*) | 1.58 (1.29 – 1.94; <0.001*) | 1.57 (1.31 – 1.88; <0.001*) | 186 / 686 (27.1%) | 1.41 (1.14 – 1.76; 0.002*) |
| Analyses adjusting in addition for any other clinical markers of disease severity | | | | | | |
| No BZRA | 1,134 / 13,695 (8.3%) | Ref. | Ref. | Ref. | 140 / 686 (20.4%) | Ref. |

| | | | | | | |
|---|---|---|---|---|---|---|
| Any BZRA | 186 / 686 (27.1%) | 1.83 (1.36 – 2.47; <0.001*) | 1.55 (1.27 – 1.90; <0.001*) | 1.54 (1.28 – 1.84; <0.001*) | 186 / 686 (27.1%) | 1.38 (1.11 – 1.72; 0.004*) |
| Analyses adjusting in addition for both respiratory depression and any other clinical markers of disease severity | | | | | | |
| No BZRA | 1,134 / 13,695 (8.3%) | Ref. | Ref. | Ref. | 136 / 686 (19.8%) | Ref. |
| Any BZRA | 186 / 686 (27.1%) | 1.83 (1.36 – 2.47; <0.001*) | 1.53 (1.25 – 1.88; <0.001*) | 1.52 (1.26 – 1.82; <0.001*) | 186 / 686 (27.1%) | 1.45 (1.16 – 1.81; 0.001*) |

* Two-sided p-value is significant (p<0.05).
Respiratory depression was defined by a respiratory rate < 12 breaths/min or a resting peripheral capillary oxygen saturation in ambient air < 90%; any other clinical markers of disease severity was defined by a temperature > 40°C or a systolic blood pressure < 100 mmHg or a respiratory rate > 24 breaths/min or a plasma lactate levels higher than 2 mmol/L.
Abbreviations: HR, hazard ratio; CI, confidence interval.

**eTable 6. Associations between individual benzodiazepine receptor agonists (BZRAs) and mortality.**

| | Number of events / Number of patients | Crude Cox regression analysis | Multivariable Cox regression analysis | Analysis weighted by inverse-probability-weighting weights | Analysis weighted by inverse-probability-weighting weights adjusted for unbalanced covariates |
|---|---|---|---|---|---|
| | N (%) | HR (95%CI; p-value) | HR (95%CI; p-value) | HR (95%CI; p-value) | HR (95%CI; p-value) |
| No BZRA | 1,134 / 13,695 (8.3) | Ref. | Ref. | Ref. | Ref. |
| Diazepam | 9 / 74 (12.2) | 1.48 (0.77 - 2.86; 0.240) | 0.85 (0.40 – 1.82; 0.678) | 0.84 (0.29 – 2.46; 0.747) | 1.20 (0.56 – 2.60; 0.636) |
| Any BZRA other than diazepam | 177 / 612 (28.9) | 3.39 (2.90 - 3.98; <0.001*) | 2.02 (1.50 – 2.73; <0.001*) | 1.63 (1.33 – 2.01; <0.001*) | 1.68 (1.40 – 2.01; <0.001*) |
| Alprazolam | 32 / 129 (24.8) | 2.56 (1.80 – 3.63; <0.001*) | 1.19 (0.82 – 1.72; 0.366) | 1.22 (0.79 – 1.89; 0.359) | 1.23 (0.82 – 1.85; 0.987) |
| Clonazepam | 9 / 43 (20.9) | 2.19 (1.14 – 4.22; 0.019*) | 1.28 (0.65 – 2.50; 0.476) | 1.32 (0.65 – 2.69; 0.442) | 2.55 (0.98 – 6.65; 0.055) |
| Midazolam | 74 / 125 (59.2) | 8.34 (6.59 – 10.57; <0.001*) | 3.72 (2.90 – 4.78; <0.001*) | 3.02 (2.29 – 3.99; <0.001*) | 3.06 (2.35 – 4.01; <0.001*) |
| Oxazepam | 57 / 234 (24.4) | 3.05 (1.97 – 4.72; <0.001*) | 1.59 (0.96 – 2.61; 0.070) | 1.64 (1.16 – 2.31; 0.005*) | 1.44 (1.06 – 1.97; 0.022*) |
| Z-drugs (zopiclone or zolpidem) | 42 / 195 (21.5) | 3.08 (1.89 – 5.03; <0.001*) | 1.79 (0.98 – 3.24; 0.057) | 1.98 (1.22 – 3.22; 0.006*) | 1.39 (0.96 – 2.00; 0.081) |
| Other BZRAs | 23 / 129 (17.8) | 1.95 (1.29 – 2.95; 0.002*) | 1.12 (0.66 – 1.90; 0.688) | 1.72 (0.92 – 3.20; 0.089) | 1.27 (0.63 – 2.54; 0.500) |
| Any benzodiazepine receptor agonist other than diazepam or midazolam | 104 / 494 (21.1) | 2.76 (1.99 – 3.83; <0.001*) | 1.60 (1.09 – 2.35; 0.018*) | 2.13 (1.52 – 2.98; <0.001*) | 1.71 (1.22 – 2.38; 0.002*) |

* Two-sided p-value is significant (p<0.05).
Abbreviations: HR, hazard ratio; CI, confidence interval.
Only individual BZRAs associated with more than 5 end-point events are presented in the table.

### eTable 7. Characteristics of patients with COVID-19 receiving diazepam versus any other benzodiazepine receptor agonist (BZRA) (N=686).

| | Diazepam (N = 74) | Other benzodiazepine receptor agonists (N = 612) | Non-exposed matched group (N=74) | Diazepam vs. Other benzodiazepine receptor agonists | Diazepam vs. Other benzodiazepine receptor agonists | Diazepam vs. Non-exposed matched group |
|---|---|---|---|---|---|---|
| | | | | **Crude analysis** | **Analysis weighted by inverse-probability-weighting weights** | **Matched analytic sample analysis** |
| | **N (%)** | **N (%)** | **N (%)** | **SMD** | **SMD** | **SMD** |
| Age | | | | 0.592 | 0.140 | 0.276 |
| *18 to 50 years* | 13 (17.6%) | 57 (9.31%) | 10 (13.5%) | | | |
| *51 to 70 years* | 34 (45.9%) | 158 (25.8%) | 27 (36.5%) | | | |
| *More than 70 years* | 27 (36.5%) | 397 (64.9%) | 37 (50.0%) | | | |
| Sex | | | | 0.458 | 0.091 | 0.114 |
| *Women* | 23 (31.1%) | 325 (53.1%) | 27 (36.5%) | | | |
| *Men* | 51 (68.9%) | 287 (46.9%) | 47 (63.5%) | | | |
| Hospital | | | | 0.253 | 0.136 | 0.073 |
| *AP-HP Centre – Paris University, Henri Mondor University Hospitals and at home hospitalization* | 26 (35.1%) | 180 (29.4%) | 28 (37.8%) | | | |
| *AP-HP Nord and Hôpitaux Universitaires Paris Seine-Saint-Denis* | 21 (28.4%) | 201 (32.8%) | 21 (28.4%) | | | |
| *AP-HP Paris Saclay University* | 17 (23.0%) | 105 (17.2%) | 15 (20.3%) | | | |
| *AP-HP Sorbonne University* | 10 (13.5%) | 126 (20.6%) | 10 (13.5%) | | | |
| Obesity [α] | | | | 0.091 | 0.024 | 0.135 |
| *Yes* | 17 (23.0%) | 118 (19.3%) | 13 (17.6%) | | | |
| *No* | 57 (77.0%) | 494 (80.7%) | 61 (82.4%) | | | |
| Smoking [β] | | | | 0.115 | 0.046 | 0.221 |
| *Yes* | 15 (20.3%) | 97 (15.8%) | 9 (12.2%) | | | |
| *No* | 59 (79.7%) | 515 (84.2%) | 65 (87.8%) | | | |
| Any medical condition [γ] | | | | 0.257 | 0.043 | 0.027 |
| *Yes* | 34 (45.9%) | 359 (58.7%) | 35 (47.3%) | | | |
| *No* | 40 (54.1%) | 253 (41.3%) | 39 (52.7%) | | | |

| | | | | | | |
|---|---|---|---|---|---|---|
| Medication according to compassionate use or as part of a clinical trial ϴ | | | | 0.012 | 0.032 | 0.064 |
| *Yes* | 18 (24.3%) | 152 (24.8%) | 16 (21.6%) | | | |
| *No* | 56 (75.7%) | 460 (75.2%) | 58 (78.4%) | | | |
| Any current psychiatric disorder ¥ | | | | 0.288 | 0.138 | 0.274 |
| *Yes* | 25 (33.8%) | 129 (21.1%) | 16 (21.6%) | | | |
| *No* | 49 (66.2%) | 483 (78.9%) | 58 (78.4%) | | | |
| Any antidepressant | | | | 0.346 | 0.046 | 0.031 |
| *Yes* | 20 (27.0%) | 265 (43.3%) | 19 (25.7%) | | | |
| *No* | 54 (73.0%) | 347 (56.7%) | 55 (74.3%) | | | |
| Any mood stabilizer medication Ω | | | | 0.129 | 0.094 | 0.096 |
| *Yes* | 19 (25.7%) | 124 (20.3%) | 16 (21.6%) | | | |
| *No* | 55 (74.3%) | 488 (79.7%) | 58 (78.4%) | | | |
| Any antipsychotic medication | | | | 0.647 | 0.220 | 0.333 |
| *Yes* | 37 (50.0%) | 126 (20.6%) | 25 (33.8%) | | | |
| *No* | 37 (50.0%) | 486 (79.4%) | 49 (66.2%) | | | |
| Number of BZRA medications | | | | 0.325 | 0.084 | 0.124 |
| *1* | 45 (60.8%) | 458 (74.8%) | 49 (66.2%) | | | |
| *2* | 22 (29.7%) | 130 (21.2%) | 18 (24.3%) | | | |
| *3 or more* | 7 (9.46%) | 24 (3.92%) | 7 (9.46%) | | | |

[α] Defined as having a body-mass index higher than 30 kg/$m^2$ or an International Statistical Classification of Diseases and Related Health Problems (ICD-10) diagnosis code for obesity (E66.0, E66.1, E66.2, E66.8, E66.9).
[β] Current Smoking status was self-reported.
[γ] Assessed using ICD-10 diagnosis codes for diabetes mellitus (E11), diseases of the circulatory system (I00-I99), diseases of the respiratory system (J00-J99), neoplasms (C00-D49), diseases of the blood and blood-forming organs and certain disorders involving the immune mechanism (D5-D8), frontotemporal dementia (G31.0), peptic ulcer (K27), diseases of liver (K70-K95), hemiplegia or paraplegia (G81-G82), acute kidney failure or chronic kidney disease (N17-N19), and HIV (B20).
[ϴ] Any medication prescribed as part of a clinical trial or according to compassionate use (e.g., hydroxychloroquine, azithromycin, remdesivir, tocilizumab, sarilumab, or dexamethasone).
[¥] Assessed using ICD-10 diagnosis codes (F00- F99).
[Ω] Included lithium or antiepileptic medications with mood stabilizing properties.
SMD>0.1 indicates substantial difference.
Abbreviation: SMD, standardized mean difference.

**eTable 8. Association between diazepam use and the endpoint of death among patients who had been hospitalized for COVID-19 and had received benzodiazepine receptor agonists at baseline (N=686).**

| | Number of events / Number of patients | Crude Cox regression analysis | Multivariable Cox regression analysis [a] | Analysis weighted by inverse-probability-weighting weights | Analysis weighted by inverse-probability-weighting weights adjusting for unbalanced covariates [b] | Number of events / Number of patients in the matched groups | Univariate Cox regression in the 1:1 ratio matched analytic sample | Univariate Cox regression in the 1:1 ratio matched analytic sample adjusting for unbalanced covariates[c] |
|---|---|---|---|---|---|---|---|---|
| | N (%) | HR (95%CI; p-value) | HR (95%CI; p-value) | HR (95%CI; p-value) | HR (95%CI; p-value) | N (%) | HR (95%CI; p-value) | HR (95%CI; p-value) |
| Diazepam | 9 / 74 (12.2) | 0.44 (0.23 – 0.86; 0.017*) | 0.49 (0.24 – 0.98; 0.044*) | 0.31 (0.13 – 0.74; 0.008*) | 0.37 (0.15 – 0.91; 0.029*) | 9 / 74 (12.2) | 0.43 (0.20 - 0.95; 0.036*) | 0.36 (0.14 – 0.96; 0.041*) |
| Other benzodiazepine receptor agonists | 177 / 612 (28.9) | Ref. | Ref. | Ref. | Ref. | 21 / 74 (28.4) | Ref. | Ref. |

* Two-sided p-value is significant (p<0.05).
[a] Adjusted for sex, age, hospital, obesity, current smoking status, any significant medical, any current psychiatric disorder, any medication prescribed according to compassionate use or as part of a clinical trial, other psychotropic medications (i.e. any antidepressant, any mood stabilizer and any antipsychotic medication), and number of BZRA medications.
[b] Adjusted for age, hospital, any current psychiatric disorder, and any antipsychotic medication.
[c] Adjusted for age, sex, obesity, smoking, any current psychiatric disorder, any antipsychotic medication, and number of BZRA medications.
Abbreviations: HR, hazard ratio; CI, confidence interval.
